## Supplementary material for "HMOX1 genetic polymorphisms and outcomes in infectious disease: a systematic review": Search strategy

("heme oxygenase" or HMOX1 or "haem oxygenase" or "heme-oxygenase" or "haem-oxygenase" or "HO-1" or “heme-oxygenase-1” ) AND

(repeat or polymorphism or variant or mutation or SNP or microsatellite or GT or genotype )
